# Promising prognostic factors in Cutaneous Leishmaniasis in regions of *Leishmaniavirus* 1 circulation

**DOI:** 10.64898/2026.09.18.26363400

**Authors:** Cipriano Ferreira da Silva-Júnior, Sayonara dos Reis, Renata Bispo Santos, Moreno Magalhães de Souza Rodrigues, Juan Miguel Villalobos Salcedo, Gabriel Eduardo Melim Ferreira, Lilian Motta Cantanhêde, Elisa Cupolillo

## Abstract

**Background:** Cutaneous leishmaniasis (CL) caused by *Leishmania (Viannia)* may involve parasite dissemination from the primary skin lesion to clinically healthy mucosal sites, a process potentially relevant to the development of mucosal leishmaniasis. Parasite burden and *Leishmania RNA Virus 1* (LRV1) have been proposed as factors influencing parasite persistence, dissemination, and treatment response. We investigated whether parasite load and LRV1 detection in skin lesions were associated with *Leishmania* in clinically healthy nasal mucosa and, in a subset of patients, with therapeutic outcome. We also assessed the relationship between LRV1 and parasite load.

**Methodology/Principal findings:** We conducted a prospective observational cohort study. Parasite load and LRV1 were assessed by qPCR in skin lesions and clinically healthy nasal mucosa from patients with localized cutaneous leishmaniasis in Rondônia, Brazilian Amazon. Samples were evaluated before treatment (D0), at the end of treatment (D20), and during follow-up (D90–180). Among 178 patients screened, CL was confirmed in 113. LRV1 was detected in skin lesions in 34.51% of patients and in nasal mucosa in 24.13%. *Leishmania* DNA was detected in clinically healthy nasal mucosa in 12.38% of patients. LRV1 detection in the skin lesion was associated with detection of *Leishmania* in the nasal mucosa. Higher parasite loads were observed in more recent lesions. Among patients evaluated for therapeutic outcome, higher parasite load before treatment was associated with treatment failure and was the factor most strongly associated with an unfavorable response in multivariate analysis.

**Conclusions/Significance:** LRV1 detection in cutaneous lesions was associated with *Leishmania* in clinically healthy nasal mucosa, although the significance of this association and its relationship with parasite dissemination require further investigation. In contrast, parasite burden was associated with therapeutic outcome, with higher pre-treatment loads observed in patients who experienced treatment failure. The absence of an association between LRV1 and parasite load suggests that these two parameters may provide distinct information regarding parasite detection at mucosal sites and therapeutic response.

**Author Summary:** Cutaneous leishmaniasis is a neglected tropical disease that mainly causes skin lesions but, in some patients, parasites may also be found at distant sites, including the nasal mucosa, even when there are no signs of mucosal disease. Understanding the factors associated with this process and with treatment response may help identify patients who require closer follow-up. We investigated whether the amount of parasites in skin lesions and the presence of *Leishmania RNA Virus 1*, a virus that naturally infects some *Leishmania* parasites, were associated with parasite detection in clinically healthy nasal mucosa and with treatment outcome in patients from the Brazilian Amazon. We found that the presence of the virus in skin lesions was associated with *Leishmania* detection in clinically healthy nasal mucosa. In contrast, patients with higher amounts of parasites in their skin lesions before treatment were more likely to experience treatment failure. These findings suggest that the virus and parasite burden may provide different information about parasite presence at distant sites and treatment response.

## INTRODUCTION

Cutaneous leishmaniasis (CL) is among the most globally significant infectious diseases and is classified by the World Health Organization as one of the top ten neglected tropical diseases. Over 12 million individuals are currently infected, and approximately 350 million people live in endemic areas. Climate change has contributed to the geographical expansion of CL, which is now endemic in 89 countries across the Americas, Europe, Africa, and Asia [1,2,3]. From 2017 to 2022, nearly 253,000 cases of CL and mucosal leishmaniasis (ML) were reported in the Americas, with the vast majority concentrated in the Andean region, Brazil, and Central America. Brazil alone accounted for over 90,000 cases during this period and continues to report the highest number of ML cases in the region [3]. Thirteen *Leishmania* species have been confirmed as etiological agents of CL in the Americas, with *L. braziliensis* most frequently associated with ML, followed by *L. guyanensis*, *L. panamensis*, and, more rarely, *L. amazonensis* [4].

In Rondônia, a highly endemic state in the western Brazilian Amazon, CL is primarily caused by *L. braziliensis*, although six other species have been identified [5]. The region has a high frequency of *Leishmania RNA Virus* 1 (LRV1), a viral endosymbiont that promotes parasite persistence, increased parasitemia, and hyperinflammatory responses that contribute to mucosal dissemination and tissue destruction in experimental infections [6]. The presence of LRV1 has also been associated with the mucosal manifestation of the human disease [7, 8], poor response to different therapeutic schemes, and increased risk of relapse, although conflicting evidence exists regarding its clinical impact [9,10,11].

ML is a complication of uncontrolled CL dissemination, which may reach mucosal sites within weeks of cutaneous infection, often before symptoms appear [12,13]. In Brazil, approximately 5% of CL cases develop mucosal lesions [14,15,16]. Early parasite dissemination to the nasal mucosa has been linked to immune alterations, including decreased IL-12 and increased IL-6 levels, as well as broader lesion distribution [17,18,19].

In disseminated CL, hematogenous or lymphatic spread in immunocompromised individuals may result in mucosal involvement reported in up to 53% of cases [20]. Nevertheless, such involvement frequently remains undetected. Furthermore, LRV1 viral load correlates with increased inflammation and tissue damage [21].

*Leishmania* has been reported in clinically healthy nasal mucosa in some endemic areas [14,22,23,24]. However, it remains unclear whether the mucosal dissemination is linked to features such as parasite load or the presence of LRV1 in the primary lesion. Higher parasite loads in primary lesions have been associated with mucosal involvement and treatment failure in experimental and clinical studies [25,26]. However, patients from Rio de Janeiro with poorer clinical outcomes paradoxically exhibited lower tissue parasitism [27]. Regarding this phenomenon, there are still few studies and limited knowledge, and a lack of methodological consistency is evident concerning the molecular techniques used to determine *Leishmania* parasite load.

Given the diversity of *Leishmania* species and the high prevalence of LRV1 in Rondônia, this setting offers a unique opportunity to explore how parasite burden and/or LRV1 presence in cutaneous lesions relate to early dissemination to the nasal mucosa and to therapeutic response to meglumine antimoniate, the first-line drug for CL in Brazil.

## MATERIAL AND METHODS

### Study design, Samples, and Clinical evaluation procedures

This prospective observational convenience-sample study was conducted according to standard care protocol of the Tropical Medicine Center of Rondônia (CEMETRON), where patient evaluation, follow-up, and sample collection were performed using a standardized Interview Form. Minimally invasive sampling of cutaneous lesions and healthy nasal mucosa was performed using a cervical brush without causing harm to the participants and according to individual clinical needs.

The study population comprised 178 patients with lesions clinically suspected of cutaneous leishmaniasis who received care at CEMETRON between February 2021 and March 2023. Samples were collected from the lesions for direct parasitological examination, and using cervical brushes, samples were obtained from both the lesion and healthy nasal mucosa for molecular analyses.

Inclusion criteria were: individuals of any age and sex, LCL confirmed by parasitological examination and/or positive qPCR, regardless of disease duration, no prior treatment and signed informed consent and assent forms when applicable. Exclusion criteria included: lack of diagnostic confirmation; previous treatment for TL; presence of mucosal lesions; primary or secondary immunosuppression; comorbidities such as cardiac arrhythmia, hepatic or renal failure; pregnancy or breastfeeding; Indigenous populations; risk of bleeding; or non-adherence to treatment protocols.

Patients underwent a structured clinical and epidemiological evaluation, lesion scarification for direct parasitological examination (178 cases), and blood was collected for routine laboratory tests. Mucosal involvement was excluded through symptom assessment and anterior examination of the nasal and oral cavities, without videopharyngolaryngoscopy. Confirmed cases were enrolled in a follow-up protocol with three scheduled evaluations: before treatment (Day-0), at the end of treatment (Day-20), and between 90 and 180 days after treatment (Day-90 and Day-180). During each visit, nasal mucosa samples were collected. Lesion samples were collected again at D-20 and D-90 only if the ulcer remained unhealed, to avoid interfering with lesion healing or causing additional trauma.

At D-0, direct microscopy and qPCR were performed for detection of Leishmania and LRV1 in cutaneous lesion and mucosa samples. Four cervical brushes were used for each patient (two per anatomical site, one for DNA and one for RNA extraction), and were stored in DNA/RNA Shield™ (Zymo Research) at −20°C until processing. All confirmed cases were referred for laboratory tests as recommended by the Brazilian Ministry of Health [28].

Treatment was conducted according to national guidelines using intravenous meglumine antimoniate at 10–20 mg/kg/day for 20 consecutive days. Clinical cure monitoring began after the treatment cycle (D-20), with assessments at D-90 to D-180. Lesions were considered clinically healed if fully re-epithelialized, with no signs of erythema, infiltration, scaling, or crusting. Persistent ulcers or inflammatory signs at D-180 were classified as unfavorable outcomes. At D-90, non-healed lesions triggered a second cycle of antimonial therapy. All follow-up visits occurred monthly for six months in accordance with national guidelines [28].

To minimize information and measurement bias, clinical assessments, sample collection, follow-up procedures, laboratory analyses, and outcome definitions were standardized throughout the study. Predefined inclusion and exclusion criteria were applied to all participants. Nevertheless, selection bias could not be completely avoided because participants were enrolled by convenience sampling and substantial loss to follow-up occurred during longitudinal assessment; these limitations were considered when interpreting treatment-outcome analyses.

### Detection of Leishmania and Leishmania RNA Virus 1 (LRV1)

#### DNA Extraction

DNA was extracted from samples using the High Pure PCR Template Preparation Kit (Roche, Germany), according to the manufacturer’s instructions. Control DNA was also extracted from *L.* (*V*.) *braziliensis* promastigotes (IOCL3621, isolate deposited at the Leishmania Collection from Oswaldo Cruz Foundation - CLIOC), previously thawed at 37°C and cultured in Schneider’s Insect Medium (Gibco) with 20% fetal bovine serum, 2% filtered human urine, and 50 μL/mL gentamicin. After three days at 24±1°C, the culture was adjusted to 1×10⁶ parasites/mL for DNA extraction using the High Pure Viral Nucleic Acid Kit (Roche, Germany).

DNA quantification was performed using a Qubit 4 fluorometer (Thermo Fisher), and purity was assessed via NanoDrop spectrophotometry. DNA was stored at −20°C.

#### PCR Targeting hsp70

Conventional PCR targeting the hsp70 gene [29] was performed on lesion samples using primers F-5’-GGACGAGATCGAGCGCATGGT-3’ and R-5’-TCCTTCGACGCCTCCTGGTTG-3’ to amplify a ∼230 bp fragment. Amplicons were sequenced (both strands) at the Fiocruz Technological Platforms Network (RPT01E/P01-003, Fiocruz, Minas Gerais, Brazil) using the ABI 3500xL Analyzer.

#### 18S qPCR for *Leishmania* Detection

Quantification of *Leishmania* DNA was performed using a multiplex qPCR targeting the 18S rRNA gene and RNase P as an endogenous control [30]. A standard curve was generated from a 1:9 mix of *L. braziliensis* (IOCL3621) DNA and commercial human DNA, serially diluted over five points (10⁶ parasites/mL to lower concentrations). Clinical samples were run alongside the standard curve. Parasite load was calculated by normalizing *Leishmania* DNA against human DNA [24,25,31].

#### RNA Extraction and qPCR for LRV1 Detection

RNA from lesion samples was extracted with the RNeasy Micro Kit (Qiagen); for nasal mucosa, the QIAamp Viral RNA Mini Kit (Qiagen) was used. cDNA was synthesized using iScript Reverse Transcription Supermix (Bio-Rad).

LRV1 was detected via SYBR Green qPCR targeting a 76 bp region using primers LRV1_ORF1_76F GACTGATTGGACGGAGGGCA and LRV1_ORF1_76R TGCTGTGGAACGTGAGGAACT [32]. Cycling conditions included 40 cycles and a melting curve analysis. Positive (*L. braziliensis* LRV1⁺) and negative controls were included.

### Data analysis

Data was summarized using medians and interquartile ranges for numerical variables, and absolute (n) and relative (%) frequencies. A Sankey diagram was used to visualize patient progression across follow-up time points. To compare parasite load over time, bootstrap estimation for repeated measures was performed [33]. Bivariate comparisons were performed using Student’s t-test for numerical variables and Fisher’s exact test for categorical variables. To assess associations between parasite load and clinical outcome (healing vs. non-healing), a generalized linear model with a binomial distribution was applied, considering parasite load, LRV1 detection in lesions and mucosa, and presence of *Leishmania* in the nasal mucosa.

### Ethical considerations

This study was approved by the Research Ethics Committee of the Tropical Medicine Center of Rondônia (CEP/CEPEM) under CAAE number 36359620.3.0000.0011. Written informed consent was obtained from all adult participants and from the parents or legal guardians of participants under 18 years of age. Written assent was also obtained from adolescents aged 12 to less than 18 years. Participation was voluntary, and participants could withdraw from the study at any time without affecting their clinical care or treatment. Confidentiality and privacy were ensured through participant coding and restricted access to study data.

## RESULTS

Between February 2021 and March 2023, 178 patients with lesions clinically suspected of cutaneous leishmaniasis (CL) were evaluated at the dermatology outpatient clinic of CEMETRON Hospital. From all patients (n=178), cutaneous lesion samples were collected for direct parasitological examination and molecular analysis. LRV1 screening in lesion samples was performed in 122 patients. Nasal mucosa samples were collected for the detection of Leishmania DNA (n=160) and LRV1 (n=88) (**Fig 1**).

**Fig 1.**
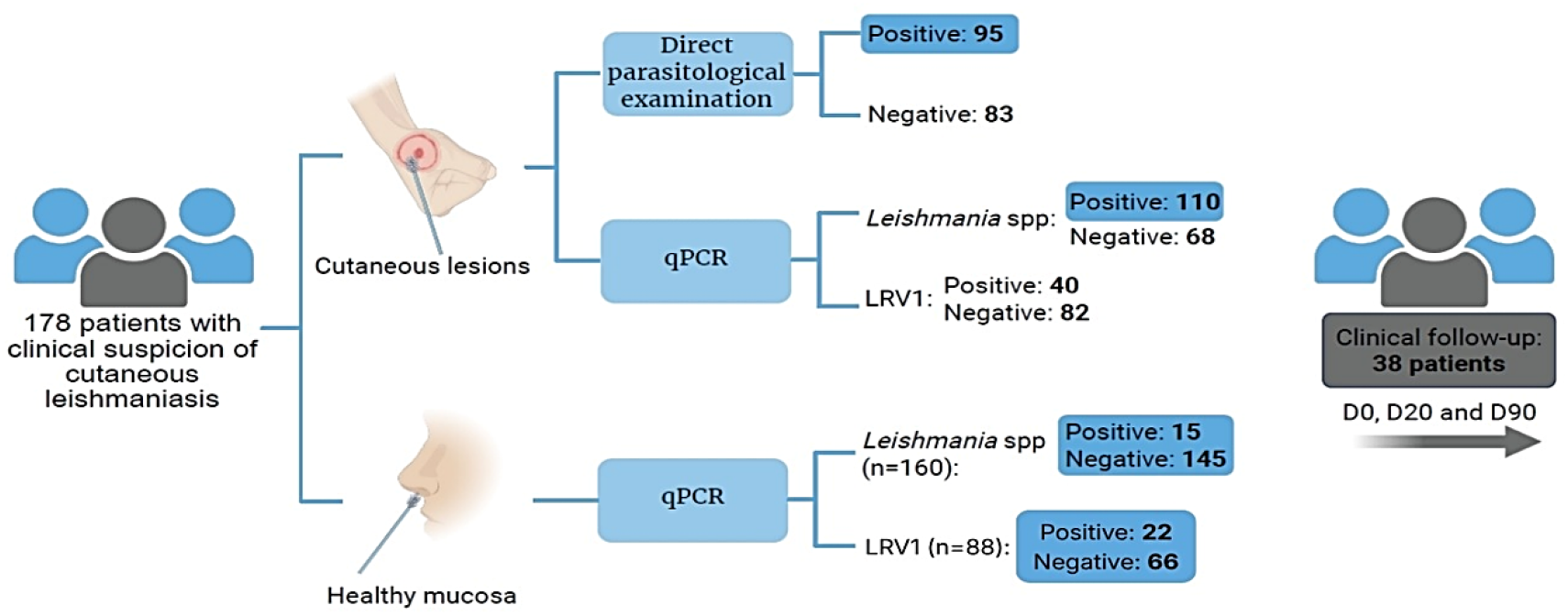
Schematic summary of the procedures performed. Results obtained for 178 patients with clinical suspicion of cutaneous leishmaniasis and no apparent involvement of the nasal mucosa.

Among the 178 patients, 95 tested positive for *Leishmania* by microscopy and 110 by qPCR: 92 (51.68%) by both, 3 (1.7%) only by microscopy and 18 (10.11%) only by qPCR (**Fig 2**). Considering a positive result in at least one of the two tests, 113 cases were confirmed and 65 were excluded.

**Fig 2.**
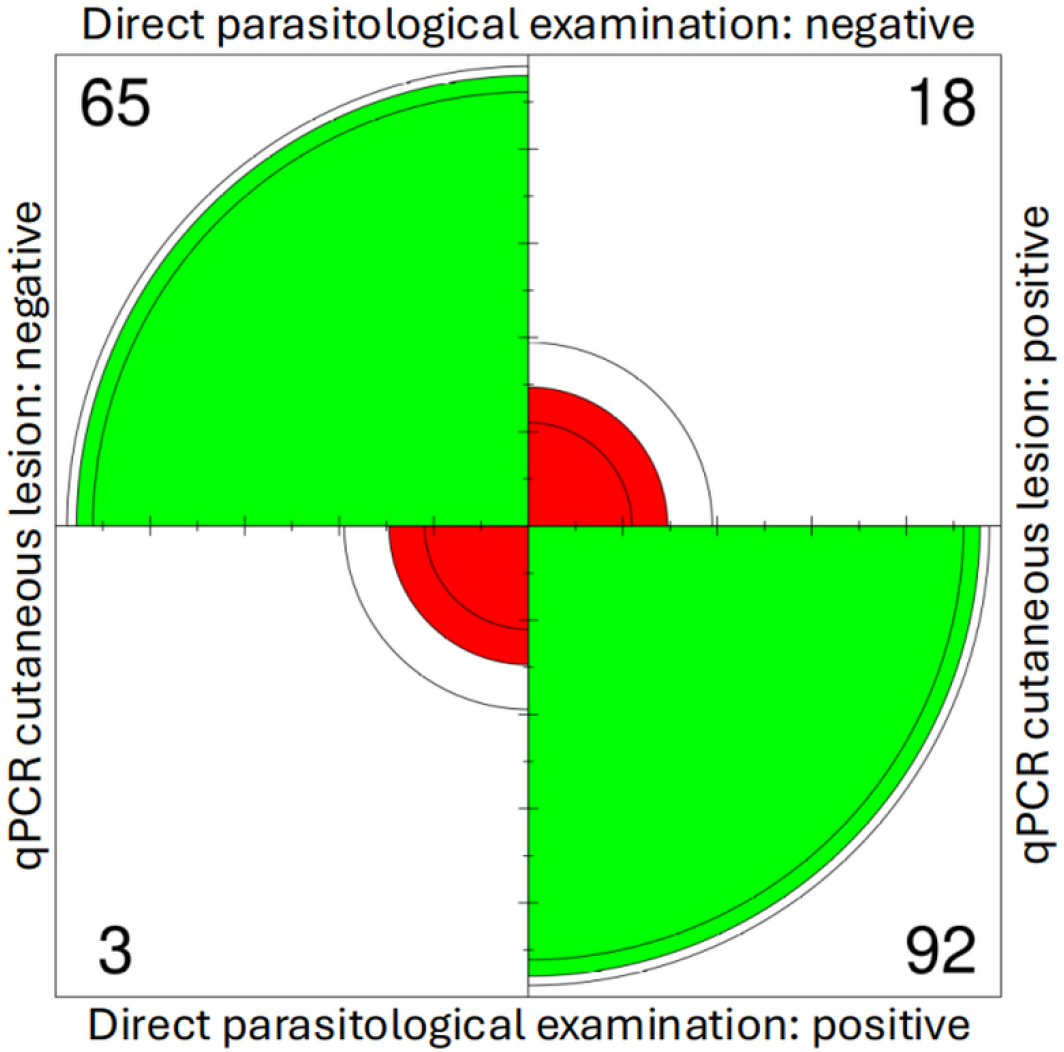
Comparative analysis of direct parasitological examination and 18S qPCR for *Leishmania* spp. Radial heatmap of the 2×2 contingency table comparing qPCR results for *Leishmania* spp. and direct parasitological examination by microscopy of the cutaneous lesion.

Among the 113 confirmed CL cases, lesion duration did not influence the diagnostic outcome (**Table S1**). Among positive cases, lesion duration ranged from under 30 days to over 10 years, with most patients (74; 67.27%) having lesions for up to 3 months. Only 15 patients (13.63%) had lesions longer than 180 days.

Finally, thirty-eight patients were followed during treatment at three time points (D-0, D-20, and D-90). After initiation of meglumine antimoniate treatment, 17 patients had healed lesions by D-20, whereas 21 still had active lesions (18 without re-epithelialization and 3 with signs of inflammation). Between D-20 and D-90, 11 of these 21 patients achieved lesion healing, resulting in a total of 28 favorable outcomes by D-90 (**Fig 3**).

**Fig 3.**
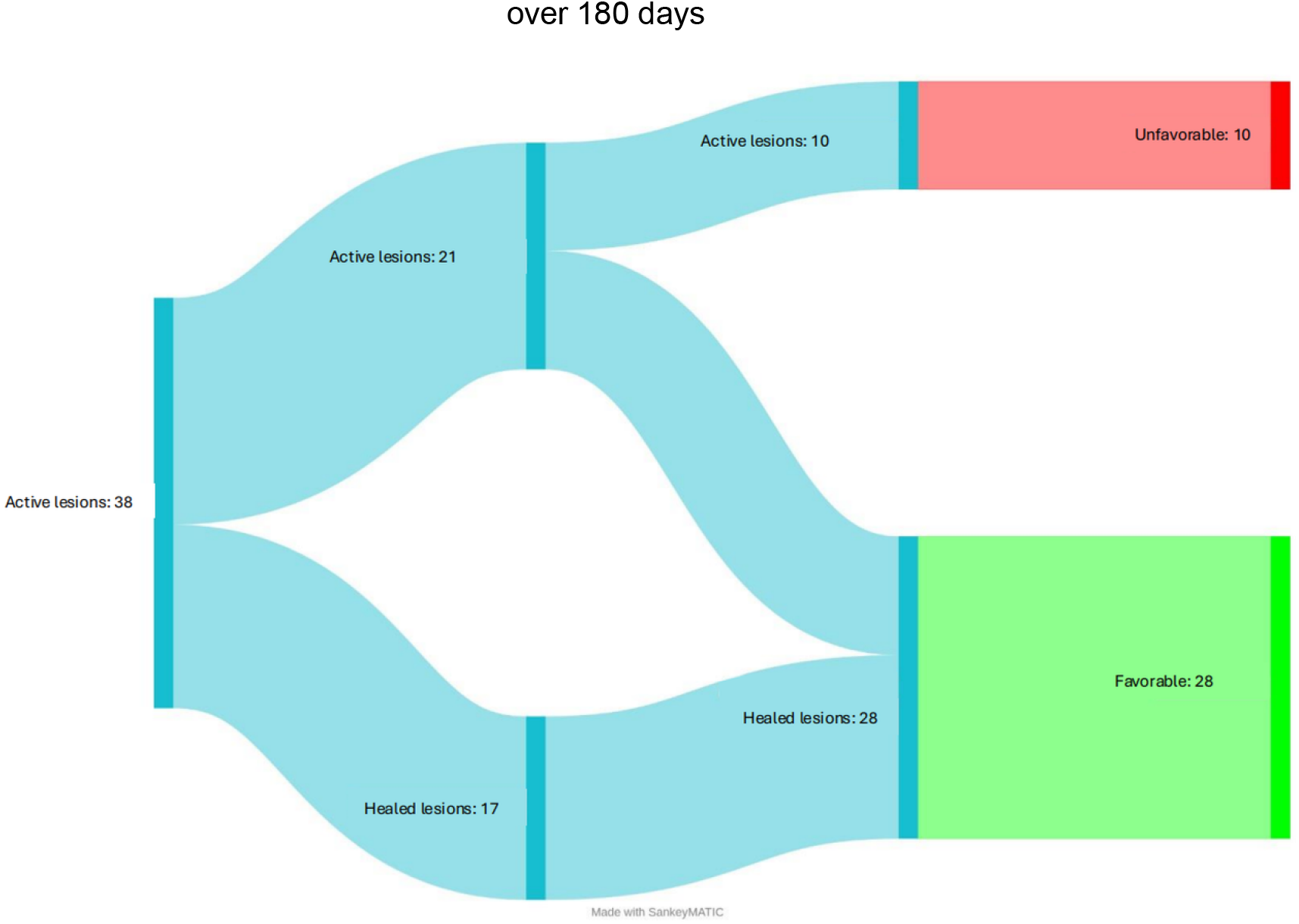
Sankey diagram illustrates the healing dynamics of lesions in 38 patients followed over 180 days.

### *Leishmania* species identified among patients with cutaneous leishmaniasis

Among the 110 patients with confirmed (qPCR) CL, 63 (57.27%) were positive by hsp70 PCR: 55 cases *L. braziliensis*, 3 *L. guyanensis*, 3 *L. lainsoni*, 1 *L. naiffi,* and 1 *L. amazonensis*. LRV1 was detected in patients infected with all identified *Leishmania* species except *L. amazonensis*, including 23 cases of *L. braziliensis*, 1 of *L. guyanensis*, 1 of *L. lainsoni*, and 1 of *L. naiffi* (**Table 1**). Among the 38 patients followed for 180 days, species identification was not achieved in 8 (21.05%) cases. Among the remaining, 24 were infected with *L. braziliensis*, 2 with *L. guyanensis*, 2 with *L. lainsoni*, 1 with *L. naiffi*, and 1 with *L. amazonensis*.

**Table 1.**
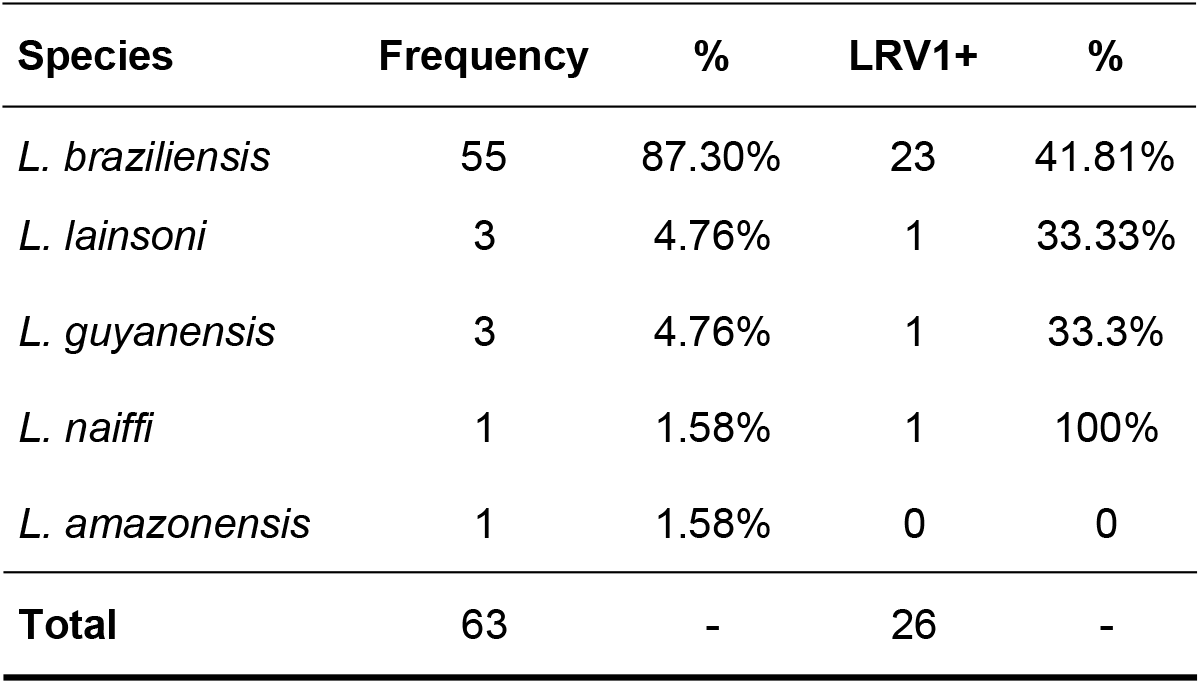
Frequency of *Leishmania* species identified by conventional PCR targeting the hsp70 gene and of LRV1 by species.

### Parasite loads (PL) of different *Leishmania* species in cutaneous lesions and in healthy nasal mucosa

The parasite load (PL) in 110 patients was 2.55 ± 1.75 (log₁₀). Among patients infected with *L. braziliensis* (n=55), the mean PL was 2.62 ± 1.69, with 34 (61.8%) presenting values above the mean. The other species (n=8) also showed parasite loads above the mean, except for one case of *L. guyanensis* with a lesion of approximately 60 days of evolution (**Table 2**).

**Table 2.**
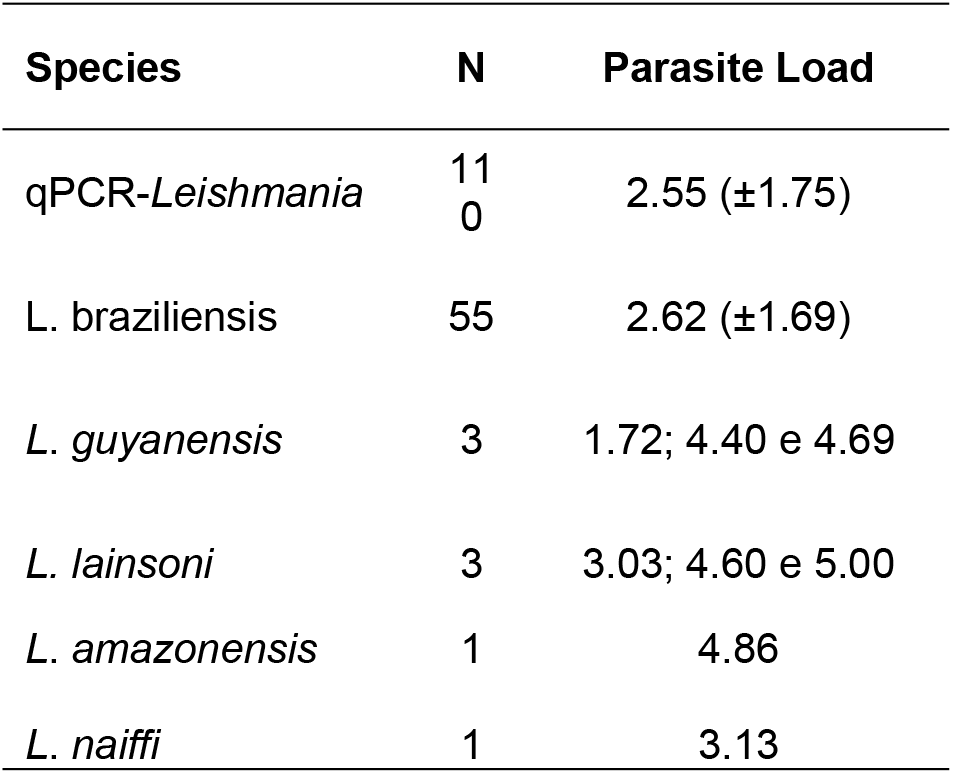
Description of parasite loads in 110 patients with confirmed CL, as determined by qPCR, and in patient groups stratified by the *Leishmania* species identified.

Among the 38 included in the follow-up, the mean lesion PL at D-0 was 3.23 ± 1.58 (log₁₀). *Leishmania* was detected in the healthy nasal mucosa of 5 patients (mean mucosal PL of 1.75 ± 2.29), four of whom presented low parasite loads. The species identified in the corresponding lesion material were *L. braziliensis* (3 cases) and *L. amazonensis* (1 case), and 3 of the 5 patients (60%) were positive for LRV1 in the lesion samples (**Table S1**).

### Percentage of patients with parasites detected in healthy nasal mucosa

In the initial mucosal screening of 88 patients for LRV1, 22 were positive. However, one patient was excluded because both *Leishmania* and LRV1 were negative in the cutaneous lesion, resulting in 21 of the 87 confirmed cutaneous leishmaniasis patients testing positive for mucosal LRV1 (24.13%). Among the 160 patients screened for *Leishmania* in the nasal mucosa, 15 tested positive, of whom 14 belonged to the group of 113 confirmed cases (12.38%). Simultaneous detection of Leishmania and LRV1 in the nasal mucosa occurred in five patients. Thus, the presence of parasites in the nasal mucosa was confirmed in 30 individuals (26.55%).

### Parasite presence in healthy nasal mucosa and its association with treatment outcome in CL patients across baseline and follow-up

At D-0, nasal mucosa samples were collected from 36 of the 38 patients included in the follow-up (two patients were excluded because D-0 samples were unavailable). Of these, 14 (38.89%) showed parasites in healthy nasal mucosa: 2 were positive for *Leishmania* only, 3 were positive for both *Leishmania* and LRV1, and 9 were positive for LRV1 only. Among these 14 patients, two had an unfavorable outcome, and 12 had a favorable outcome. Among 9 patients with unfavorable outcome (group A), 2 (22.22%) had parasites in the mucosa. Among the 27 with a favorable outcome (group B), 12 (44.44%) had parasites in the mucosa, a paradoxically higher frequency that was not statistically significant (p= 0.429795). Considering all follow-up time points, 25 (65.79%) patients had parasites in the mucosa: 19 (67.85%) in the favorable group and 6 (60%) in the unfavorable group, with no significant difference (p = 0.9511). Four patients were positive for *Leishmania* in the mucosa at D-90/D-180. Three (75%) had an unfavorable outcome. One patient presented positive mucosal qPCR and negative qPCR in the cutaneous lesion at the same point, evolved with an unfavorable outcome one had a favorable outcome, with complete re-epithelialization of the ulcer noted at D-90, although some degree of infiltration remained at the scar margins, which disappeared over the 180-day follow-up. Cases such as this may require prolonged observation periods, up to 180 days, for cure confirmation.

### Relationship between *Leishmania* parasite loads in cutaneous lesions and presence of parasites in healthy nasal mucosa

Among the 21 patients with LRV1 in healthy nasal mucosa at D-0, the mean lesion was PL 2.68 (SD = 1.77), which did not differ from the overall mean of 2.55 ± 1.75(t = 0.318, p = 0.75). Eleven of the 21 patients had PL values above the mean.

*Leishmania* was detected in the nasal mucosa of 14 patients (12.38%; 14/113). Ten of these had PL in the lesion above the overall mean, and four below (mean = 2.55; SD = 1.75).

The 30 patients presented *Leishmania* and/or LRV1 in the nasal mucosa (5 had both, 9 had *Leishmania* only, and 16 had LRV1 only), had a mean lesion PL of 2.94 ± 1.48, not differing from mucosa-negative patients (t=0.86, p=0.38) (**Fig 4**). No association was found between lesion PL and mucosal parasites (t=0.86, p=0.38) (**Fig 5**).

**Fig 4.**
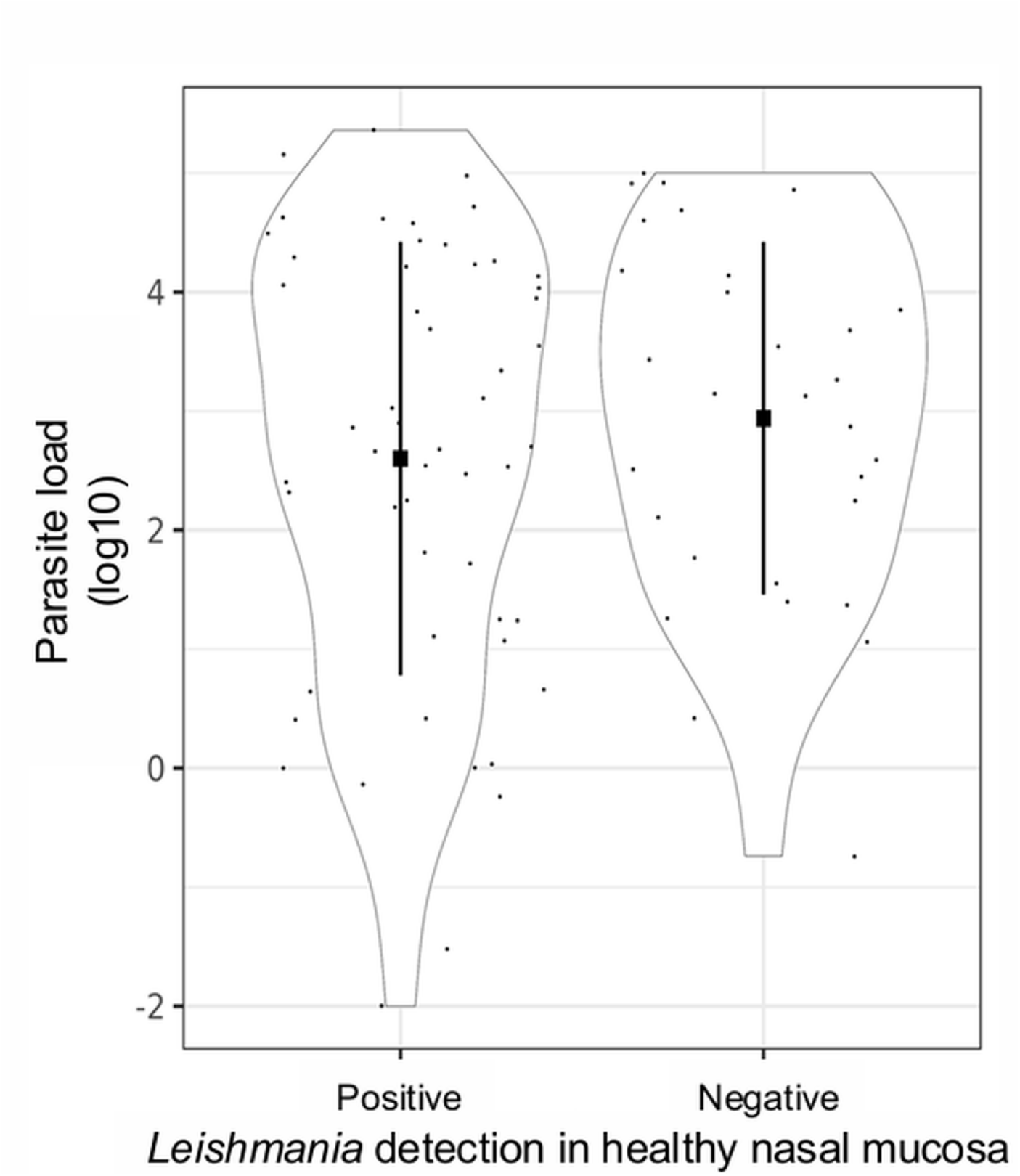
Violin plot showing the relationship between *Leishmania* spp. parasite load in the cutaneous lesion of 105 patients and the presence of the parasite (*Leishmania*, LRV1, or both) in healthy nasal mucosa. Dots represent observed values; squares indicate the mean, and the solid black line represents the mean ± standard deviation. t = 0.86, p = 0.39.

**Fig 5.**
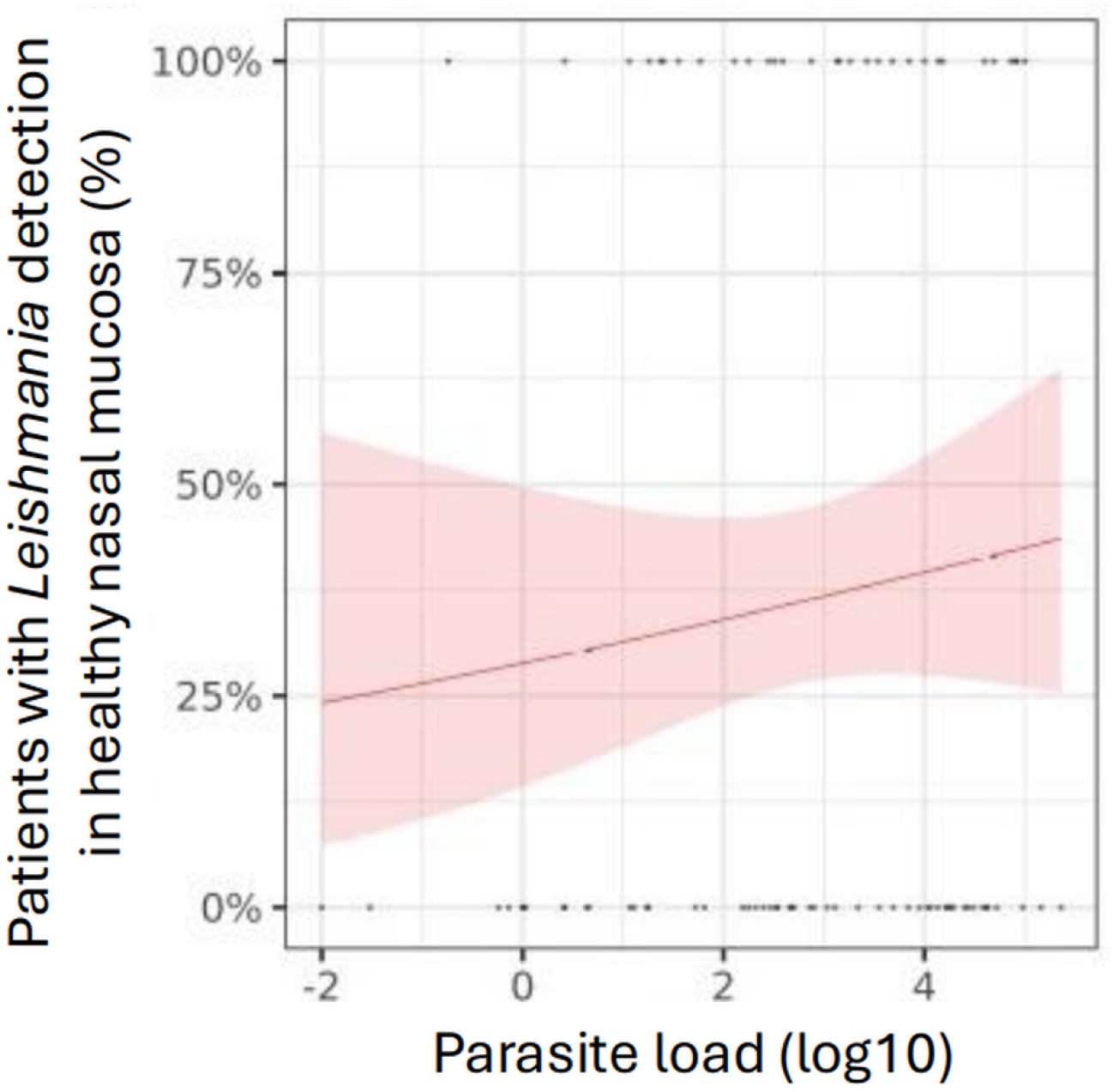
Percentage of patients with parasite presence in the nasal mucosa in relation to *Leishmania* parasite load in the cutaneous lesion. The red line indicates the mean value from the model. The pink area represents the 95% confidence interval around the mean. Dots correspond to the observed results for *Leishmania* and/or *Leishmania RNA virus 1*. In this case, points at y = 0% indicate negative patients, while y = 100% indicate positive patients.

### Correlation between *Leishmania* parasite load in lesions and duration of cutaneous disease

Parasite load (PL) decreased with increasing disease duration. Patients with lesions of ≤90 days duration (Group A) exhibited significantly higher parasite loads (2.85 ± 1.53) than those with lesions of >90 days duration (Group B; 2.06 ± 1.92) (t = 2.133, p = 0.035). This suggests a relationship between lesion evolution time and parasite load (**Fig 6**).

**Fig 6.**
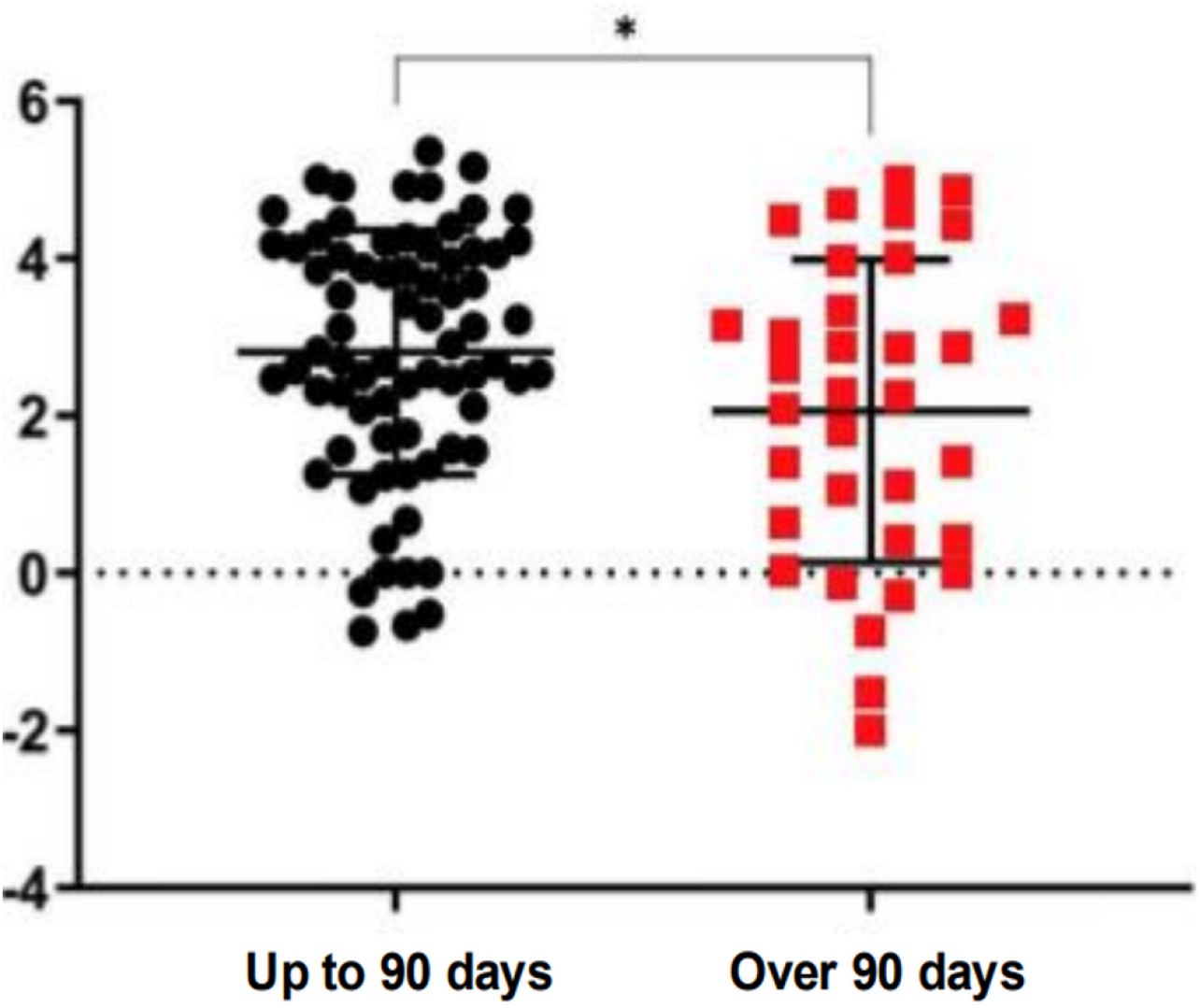
Distribution of parasite load in Log10 as a function of disease duration. Patients were grouped according to their reported lesion duration: one group reported less than 90 days (black circles), and the other reported lesion duration longer than 90 days (red squares). t = 2.133, *p = 0.03*

### Correlation between *Leishmania* parasite loads in cutaneous lesions before and after treatment with Meglumine Antimoniate

Among the 38 patients included in the follow-up, 28 had favorable outcomes and 10 had unfavorable outcomes. Mean lesion parasite load (PL) decreased from 3.23 ± 1.58 at baseline (D-0) to 2.18 ± 1.27 at the end of treatment (D-20), followed by a slight increase to 2.46 ± 1.37 at D-90/D-180 (**fig 7A**). Patients who experienced an unfavorable outcome had a significantly higher mean baseline parasite load (log10) at D-0 (mean = 4.44; SD = 0.66) compared to those who were cured (mean = 2.78; SD = 0.66; p < 0.05) (**Fig 7B**).

**Fig 7.**
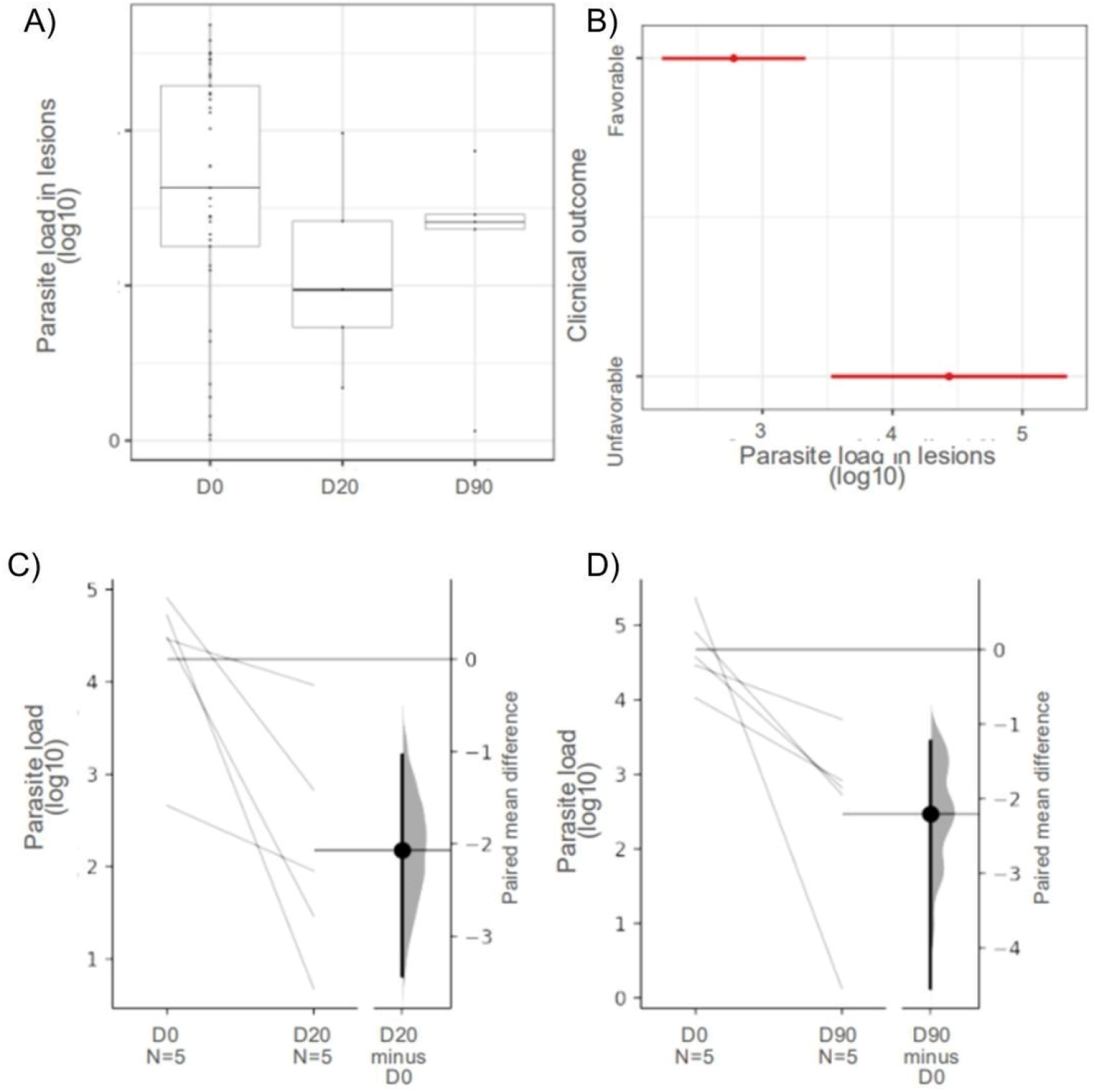
Correlation of *Leishmania* spp. parasite load in cutaneous lesions over time and according to clinical outcome. **(A)** Boxplot of overall parasite load throughout patient follow-up. **(B)** Comparison of parasite load at D-0 between patients who progressed to cure and those with an unfavorable outcome. Paired differences between **(C)** D-0 and D-20, and **(D)** D-0 and D-90 / D-180 are shown in the Gardner–Altman plots above. In each plot, groups are displayed on the left panel as a slope graph: each observation is connected to its pair by a line. The values obtained in 5,000 bootstrap resamples are shown in the right panel as a density plot, with a vertical line at the null value (0).

Paired analysis comparing D-0 and D-90 / D-180 showed a significant reduction in lesion PL during follow-up (−2.2; 95% CI: −4.4, −1.22; p < 0.05) (**Fig 7D**). It was not possible to compare D-20 and D-90/D-180 because only two patients had paired samples at those time points.

In the multivariate model using baseline (D-0) data, only lesion PL was associated with the therapeutic outcome (OR =5.63; 95% CI: 1.73—47.25) (**Table 3**). A similar result was observed in a model that considered all three time points (i.e., D-0, D-20, and D-90). In this model, an increase in the average PL within the same individual was associated with an average increase in OR of 2.66 (95% CI: 1.29–7.65) (**Table 3**).

**Table 3.** Result of generalized linear model fitted to a binomial probability distribution.

| Variable | Clinical outcome baseline |  |  | Final treatment outcome |  |  |
| --- | --- | --- | --- | --- | --- | --- |
|  | OR | 95%CI | p-value | OR | 95%CI | p-value |
| Parasite load in CL | 5.63 | 1.73 – 47.25 | 0.03 | 2.66 | 1.29 – 7.65 | 0.02 |
| LRV1 in CL | 5.39 | 0.65 – 71.70 | 0.14 | 1.62 | 0.30 – 9.44 | 0.57 |
| *Leishmania* in nasal mucosa | 0.18 | 0.01 – 1.64 | 0.16 | 0.52 | 0.08 – 3.00 | 0.45 |
| N observations | 35 |  |  | 35 |  |  |
| Adjusted R<sup>2</sup> | 0.407 |  |  | 0.211 |  |  |

### Frequency of LRV1 in cutaneous lesions and parasite load of patients

Among the 122 patients screened at D-0, LRV1 was detected in 40 patients (32.78%). Among the 113 confirmed CL diagnoses at D-0, 39 (34.51%) were positive for LRV1, of whom 8 (20.51%) were negative by direct microscopy. LRV1-positive lesions exhibited a higher mean PL of 2.86 (SD = 1.85) than LRV1-negative lesions (2.37; SD = 1.52), however, this difference was not statistically significant (t=1.43, p = 0.155) (**Fig 8A**). No significant association was observed between the presence of LRV1 and lesion PL (t = 1.41; p = 0.15, **Fig 8B**).

**Fig 8.**
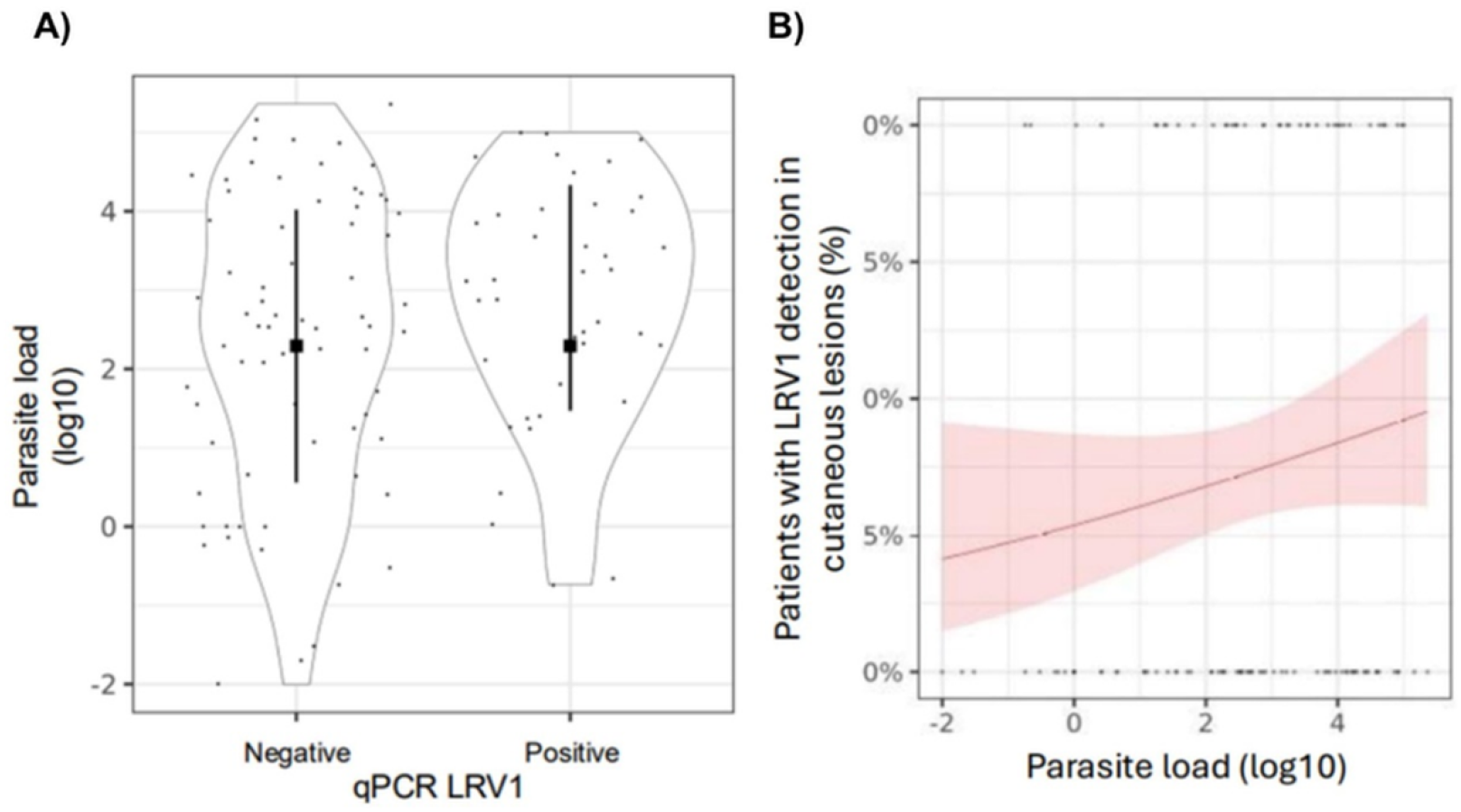
Relationship between Leishmania spp. Parasite load in cutaneous lesions (n = 110) and LRV1 presence, showing distribution of parasite load and proportion of LRV1-positive cases according to lesion parasite burden. **A)** Violin plot showing the relationship between *Leishmania* spp. parasite load in the cutaneous lesion of 110 patients and the presence of LRV1 in the same lesion. Dots represent observed values; squares indicate the mean, and the solid black line represents the mean ± standard deviation. t = 1.43, p = 0.155. **B)** Percentage of patients with LRV1 in the cutaneous lesion in relation to *Leishmania* parasite load in the same lesion. The red line indicates the mean value from the model. The pink area represents the 95% confidence interval around the mean. Dots correspond to the observed results for LRV1. In this case, points at y = 0% indicate negative patients, while y = 100% indicate positive patients.

### Relationship between LRV1 positivity in cutaneous lesions and the presence of parasites in healthy nasal mucosa

Patients positive for LRV1 in the lesion had, on average, a 4.1-fold higher chance (95% CI: 1.60–10.5; z = 2.90; p < 0.05) of presenting *Leishmania* in the nasal mucosa compared to LRV1-negative patients (**Fig 9**). Among the 140 LRV1-positive patients, 19 (47.5%) had parasites in the nasal mucosa, of whom 12 (63.16%) had PL in the cutaneous lesion above the mean. In contrast, among the 82 LRV1-negative patients, only 10 (12.19%) had parasites detected in the nasal mucosa, and most of them (60%) had lesion PL below the mean.

**Fig 9.**
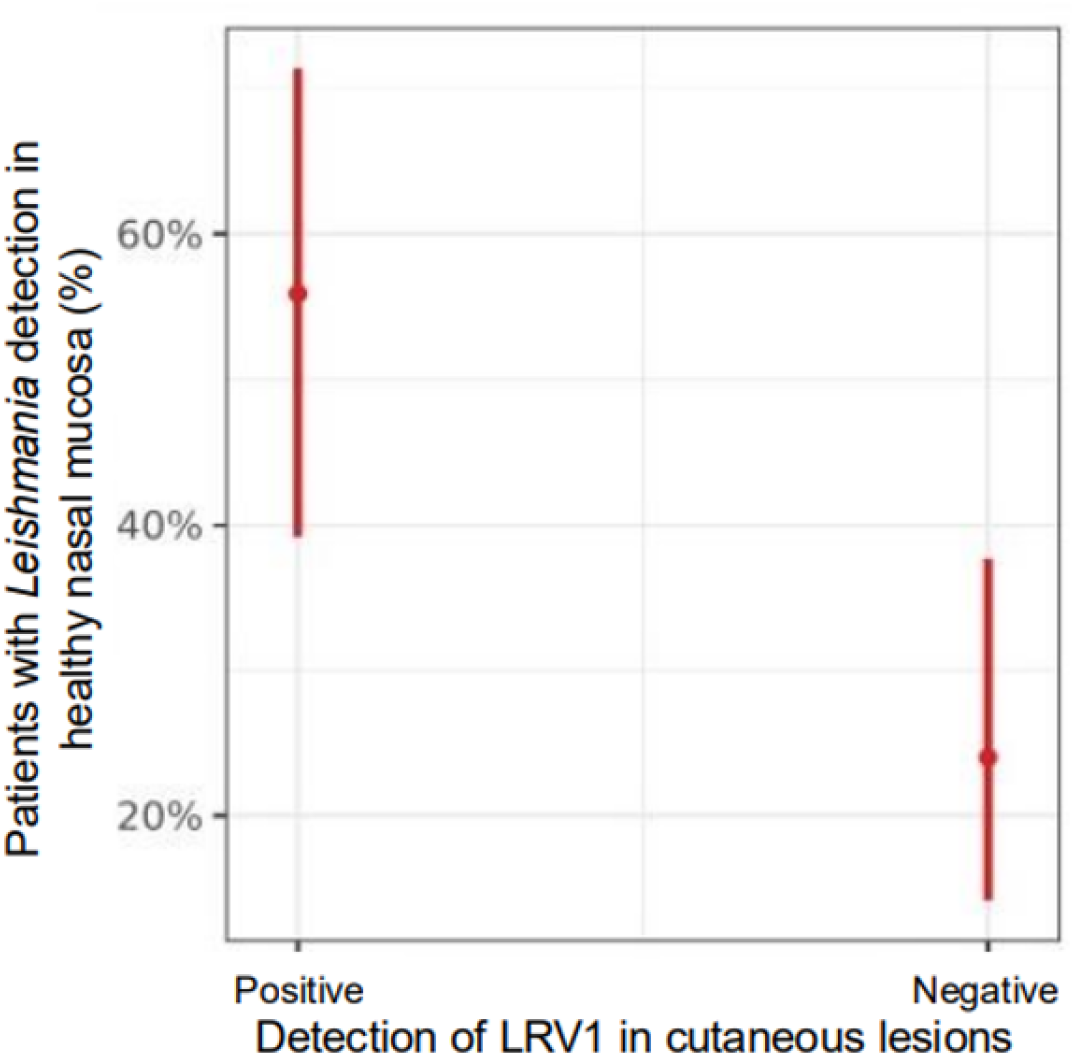
Relationship between LRV1 positivity in the cutaneous lesion and the presence of parasites in the nasal mucosa.

### Correlation between the *Leishmania*-LRV1 endosymbiosis in cutaneous or mucosal lesions and therapeutic failure

Among the 38 patients, LRV1 was detected in 16 patients (42.10%). Of these, 11 (11/28; 39.30%) had favorable outcomes and 5 (5/10; 50%) unfavorable outcomes, while among the 22 patients without LRV1, 17 had favorable outcomes and 5 unfavorable outcomes.

At D-20, among the 18 patients who still had open lesions, the same analysis was performed in 17 individuals. A reduction in positivity was observed, with LRV1 detected in only 5 patients (29.41%). Among those, four evolved with a favorable outcome (cure) and one with an unfavorable outcome.

At D-90 / D-180, among 9 patients evaluated (all with unfavorable outcomes), LRV1 was not detected in the cutaneous lesion. In one patient from the unfavorable outcome group, no sample was collected at D-90 / D-180 because the lesion had re-epithelialized, although it remained highly infiltrated. This same patient developed a new ulceration between D-90 and the final observation time point (180 days) (**Table 4**).

**Table 4.** Description of the main results of the tests performed on patients throughout the study.

|  | Favorable clinical outcome (n=28) |  |  | Unfavorable clinical outcome (n=10) |  |  |
| --- | --- | --- | --- | --- | --- | --- |
|  | D-0 | D-20 | D-90 | D-0 | D-20 | D-90 |
| Parasite load in CL | 2.78 (1.59) | 1.95 (.) | - | 4.44 (0.66) | 2.24 (1.46) | 2.46 (1.37) |
| LRV1 in CL |  |  |  |  |  |  |
| Negative | 17 (60.71%) | 7 (63.63%) | 0 | 5 (50.00%) | 5 (83.33%) | 9 (100%) |
| Positive | 11 (39.29%) | 4 (36.37%) | 0 | 5 (50.00%) | 1 (16.67%) |  |
| LRV1 in nasal mucosa |  |  |  |  |  |  |
| Negative | 19 (67.86%) | 13 (59.09%) | 18 (64.28%) | 6 (75.00%) | 7 (87.50%) | 9 (90.00%) |
| Positive | 9 (32.14%) | 9 (40.91%) | 10 (35.72%) | 2 (25.00%) | 1 (12.50%) | 1 (10.00%) |
| *Leishmania* in nasal mucosa |  |  |  |  |  |  |
| Negative | 23 (82.14%) | 26 (100%) | 27 (96.40%) | 9 (100%) | 8 (100%) | 7 (70.00%) |
| Positive | 5 (17.86%) | 0 | 1 (3.60%) | 0 | 0 | 3 (30.00%) |

At D-0, 32.1% (9/28) of the patients who were cured showed LRV1 in the mucosa, whereas among those with an unfavorable outcome, the proportion was 25% (2/8). At D-20, 40.9% (9/22) of those who were cured had positive LRV1 results in the mucosa, compared to only 12.5% (1/8) among those with unfavorable outcomes. At D-90 / D-180, 35.7% (10/28) of the cured patients and 10% (1/10) of those with unfavorable outcomes were positive for LRV1 in the mucosa.

## Discussion

Diagnostic performance reinforces the added value of molecular tools for CL confirmation. Direct parasitological examination identified the majority of cases (84.07%), but qPCR demonstrated higher sensitivity (97.34%), capturing patients missed by microscopy. Discordant results highlight the limitations of conventional methods in low parasite burden or heterogeneous lesions [34–37].

This study followed 113 patients with confirmed CL to evaluate prognostic factors associated with treatment outcomes using meglumine antimoniate, including only first-time, untreated infections. Of these, 38 were monitored for 180 days, with loss to follow-up reflecting geographic and economic barriers, and reduced services during the COVID-19 pandemic. The therapeutic failure rate following first-line treatment with meglumine antimoniate, after a 6-month observational follow-up, was 26.31%, consistent with previous reports, including 21.9% in Peru, reaching 30.4% among cases caused by *L. braziliensis* [38, 39]. In contrast, studies conducted in Bolivia and Brazil reported lower failure rates of 7% and 16%, respectively [39,40]. The variability likely reflects differences in study design, endemic settings, therapeutic regimens, and the diversity of infecting *Leishmania* species. In the absence of robust regional data, the relatively high failure rate observed in our study may be partially explained by follow-up bias as patients with unfavorable outcomes were more likely to return for evaluation, resulting in the underrepresentation of patients who achieved early clinical cure. The relatively small number of participants completing longitudinal follow-up also resulted in limited statistical precision, as reflected by the confidence intervals of some effect estimates. Therefore, although the observed associations indicate a consistent direction of effect, their magnitude should be interpreted cautiously. Moreover, because patients with unfavorable outcomes were more likely to remain under follow-up, the observed treatment failure rate and the strength of the association between parasite load and unfavorable outcome may have been overestimated. However, the magnitude of this potential bias cannot be quantified because outcome information was unavailable for participants lost to follow-up.

The positivity rate of direct parasitological examination was slightly higher than reported in previous studies [41, 42, 43], likely reflect technical expertise and standardized laboratory procedures. Most cases (67.27%) had lesions <3 months, associated with higher parasite burden and easier microscopic detection [25]. Accordingly, PL was higher in lesions ≤90 days. Diagnosis of lesions >6 months remains challenging due to reduced parasite density [44]. Regarding the positivity of *Leishmania* detection in lesions by qPCR (97.34%), the observed rate is consistent with those reported in the literature [30,36,45]. Three patients were positive only by direct microscopy, an uncommon discrepancy that highlights the complementary value of molecular and conventional diagnostic methods. This finding likely reflects sampling variability, as cervical brush specimens may include non-infected areas due to the heterogeneous distribution of amastigotes within lesions, resulting in false-negative molecular results [36, 40].

Conventional PCR confirmed the predominance of *L. braziliensis* as the main etiological agent of CL in the Rondônia region. However, the circulation of other species was also observed, including *L. guyanensis*, *L. lainsoni*, and *L. amazonensis* [6, 46]. This study represents the first report of *L. naiffi* in patients treated at CEMETRON. Although novel in this clinical setting, this finding was not unexpected, as this species has previously been detected in phlebotomine sand flies in Rondônia [47], and a case of CL in a patient from Manaus who reported acquiring the infection in Rondônia [48]. The diversity of *Leishmania* species identified in Rondônia highlights the ecological complexity of the Amazon region. Nevertheless, species-level identification was not achieved for all patients, underscoring the methodological limitations of the molecular approach employed. The mean PL among the 38 patients who were longitudinally followed was higher than that observed in the initial cohort, likely reflecting selection bias toward patients with unfavorable outcomes and higher parasite burdens. Although *L. braziliensis* infections are typically associated with low parasitism [49], a modest increase in parasite load was observed in cases caused by this species. Notably, 43.63% of these patients belonged to the follow-up cohort, and regional genetic variability within *L*. *braziliensis* populations may have contributed to the distinct profiles observed in Rondônia [50–53]. The remaining species exhibited higher parasite loads, suggesting greater tissue parasitism [54, 55], although the number of cases was limited.

*Leishmania* and the PL in the nasal mucosa were detected in only five patients, most of whom exhibited low mucosal PL at D-0. This finding is consistent with previous evidence indicating that mucosal parasite burden is generally lower than that observed in cutaneous lesions and tends to increase only in more severe forms of the disease [25, 49].

The detection of LRV1 in 60% of the cohort patients supports the findings that the presence of LRV1 in cutaneous lesions was significantly associated with the detection of *Leishmania* in the mucosa. The species identified in cutaneous lesions included *L. braziliensis* and *L. amazonensis*. As species identification was not performed on nasal mucosa samples, it cannot be assumed that the parasite detected at this site corresponded to *L*. *amazonensis*. This is particularly relevant given that LRV1 is an endosymbiont predominantly associated with parasites of the *Viannia* subgenus; its detection in nasal mucosa samples at D20 and D90 from a patient with *L. amazonensis* in the cutaneous lesion suggests a possible mixed infection involving a *Viannia* species. Heterogeneous parasite subpopulations, polyclonal inocula [53], and independent multiple infections support this plausibility [52]. Parasite persistence in lesions at D-20/D-90 was more frequent in patients with unfavorable outcomes, consistent with *L. braziliensis* chronicity, latency, and metastatic potential [52] and its predominance (90%) among cases of treatment failure in this cohort.

The prevalence of *Leishmania* DNA in healthy nasal mucosa was higher than the 7.8% reported in a non-Amazonian region of Brazil [22], but lower than the 45-61.5% reported in Colombia among patients with acute cutaneous leishmaniasis without clinical evidence of mucosal involvement [23, 56], where the mucosal tropism of *L. panamensis* is well documented [57].

The detection of LRV1 in the mucosa exceeded that of the *Leishmania* DNA target at the same anatomical site. In absolute terms, fourteen patients who tested positive for LRV1 in the mucosa were negative for *Leishmania* by molecular assays, mirroring the pattern observed in cutaneous ulcers and suggesting greater analytical sensitivity for LRV1 detection. The proportional detection of LRV1 was higher in cutaneous ulcers than in nasal mucosa, consistent with the cutaneous lesion representing the primary site of infection, followed by hematogenous and lymphatic dissemination. In cases of concomitant ML, dissemination to mucosal tissues occurs early and simultaneously with the onset of the cutaneous lesion, whereas in late ML, mucosal involvement develops at a later stage [14].

Even considering that the presence of LRV1 may indicate concomitant *Leishmania* infection in the nasal mucosa, the detection rate of parasites in asymptomatic mucosa remains substantially lower than that reported in symptomatic ML cases (70.3%) in the region [8]. This reinforces that the mere presence of *Leishmania* in the nasal mucosa does not necessarily correlate with the development of clinically apparent mucosal disease.

Although not statistically significant, patients with TL who presented parasites (*Leishmania*, LRV1, or both) in the healthy nasal mucosa tended to exhibit higher PL. The detection of parasites in asymptomatic mucosa suggests a strain profile with greater capacity for persistence, dissemination, and possible therapeutic refractoriness, potentially associated with LRV1 coinfection [6, 58–60], consistent with lineages able to escape the primary cutaneous site and colonize other tissues, including mucosal surfaces [23, 56, 57]. Conversely, in scenarios of sustained clinical cure, the detection of *Leishmania* in mucosal tissue is rare and is generally associated with cases of active ML. It is important to emphasize that mucosal dissemination is not determined solely by parasite burden but rather involves multiple parasite- and host-related factors.

Parasite load declined with increasing disease duration, with lesions of <90 days exhibiting significantly higher parasite loads than older lesions (>90 days). This finding is consistent with the inverse relationship between PL and disease duration previously reported by Jara et al [25], as well as with the greater diagnostic difficulty associated with lesions persisting for more than six months [45], likely reflecting the reduction in tissue parasitism accompanying the establishment of the inflammatory response [61, 62].

Among the 38 patients followed, a reduction in PL was observed during treatment with meglumine antimoniate, followed by a slight increase after treatment completion, although still below baseline levels. This pattern suggests the absence of sterile cure and possible parasite persistence. The decrease in PL is also influenced by the natural course of the disease [63, 64].

In the literature, very early treatment has been associated with treatment failure [65, 66], however, this was not observed in this cohort. In contrast, patients with unfavorable outcomes presented significantly higher baseline PL (p < 0.05), indicating the prognostic value of pre-treatment parasite burden. Multivariate models confirmed this association, showing that an increase in PL at D0 substantially raises the risk of an unfavorable outcome (OR ≈ 5.63). Among the 10 unfavorable, high-PL cases, nine were *L. braziliensis* and one *L. guyanensis*; six non-*braziliensis* cases with high PL did not fail, though this cannot be confirmed statistically.

In this study, the presence of LRV1 in cutaneous lesions was not associated with parasite load, but was significantly linked to parasite detection in the nasal mucosa, increasing this likelihood by 4.1-fold. This supports a potential role for LRV1 in parasite dissemination. While species such as *L. braziliensis* are inherently associated with mucosal involvement, LRV1 has been proposed as a cofactor for disease severity, although evidence remains inconsistent [6, 58, 61, 67]. No significant association was observed between LRV1 and therapeutic failure following treatment with meglumine antimoniate, in line with the heterogeneous findings reported in the literature [10, 9]. Overall, LRV1 may contribute to dissemination and severity, but does not appear to independently determine clinical outcomes.

## Concluding remarks

Although the presence of LRV1 in cutaneous lesions was associated with parasite positivity in healthy nasal mucosa, the 180-day follow-up was insufficient to determine its long-term significance. Extended follow-up may clarify its role in late-onset ML, particularly because some patients with high parasite loads had favorable outcomes, while others showed persistent lesions despite the absence of detectable parasites, suggesting that additional factors, including parasite quiescence, may influence treatment response.

In conclusion, this study highlights the diagnostic and prognostic value of qPCR for sensitive detection and quantification of parasite load in TL. Although higher baseline parasite load was associated with unfavorable outcomes, neither mucosal dissemination nor LRV1 alone determined clinical evolution. These findings reinforce that TL outcome is multifactorial and support integrated molecular diagnostics with long-term follow-up to improve prognostic assessment.

## Author contributions

Conceptualization: Lilian Motta Cantanhêde and Elisa Cupolillo; Methodology: Sayonara dos Reis, Renata Bispo Santos; Formal analysis and investigation: Cipriano Ferreira da Silva-Júnior, Sayonara dos Reis, Renata Bispo Santos, Moreno Magalhães de Souza Rodrigues; Writing - original draft preparation: Cipriano Ferreira da Silva-Júnior, Lilian Motta Cantanhêde and Elisa Cupolillo; Writing - review and editing: Cipriano Ferreira da Silva-Júnior, Sayonara dos Reis, Renata Bispo Santos, Moreno Magalhães de Souza Rodrigues, Gabriel Eduardo Melim Ferreira, Lilian Motta Cantanhêde and Elisa Cupolillo; Funding acquisition: Gabriel Eduardo Melim Ferreira and Elisa Cupolillo; Supervision: Juan Miguel Villalobos Salcedo and Elisa Cupolillo

## Acknowledgments

We sincerely appreciate Claudino Limeira de Sousa, a professional who carried out the collection of samples and performed direct parasitological diagnostic by microscopic examination and Fiocruz Technological Platforms Network (RPT01E/P01-003, Fiocruz, Minas Gerais, Brazil) for performing the sequencing of the clinical samples.

## Data Availability

The individual-level data underlying the findings of this study are provided in **Table S1**. Additional study data cannot be made publicly available due to privacy and ethical restrictions concerning human participants. Requests for access to additional data may be considered by the corresponding author and are subject to approval by the relevant ethics committee.

## Supporting Information

**Table S1.** Description of the results obtained for the 178 patients with leishmaniasis included in the study, highlighting the 38 patients who were followed until the end of treatment. **Legend:** NP = not performed; NI = Not informed; rows in gray represent the 38 patients followed throughout the study.

**Table S2.** STROBE statement checklist.

